# Viral Cultures for Assessing Airborne Transmission of SARs-CoV-2: a Systematic Review Protocol (version 1)

**DOI:** 10.1101/2022.01.28.22270021

**Authors:** Carl J. Heneghan, Elizabeth A. Spencer, Jon Brassey, Elena C. Rosca, Annette Plüddemann, David H. Evans, John M. Conly, Susanna Maltoni, Tom Jefferson, Igho J. Onakpoya

## Abstract

This is a protocol for a systematic review that aims to evaluate the role of viral cultures for assessing airborne transmission of SARS-CoV-2. The review will address the following research questions:

Are airborne samples infectious?

If so, what proportion are infectious, and what is the distance and duration of infectiousness in the air?

What is the relationship between infectiousness and airborne PCR cycle threshold (Ct)?

Is there evidence of a chain of transmission that establishes an actual instance of airborne transmission of SARS-CoV-2?

What circumstances might facilitate infectious viruses being airborne over long distances?

We will search LitCovid, medRxiv, Google Scholar, and the WHO Covid-19 database to identify relevant studies. We will include studies reporting airborne transmission attempting viral culture or serial qRT-PCR with or without genomic sequencing. Predictive or modelling studies will be excluded. We will assess the quality of included studies using previously published criteria.

## Background

Airborne transmission of an infectious agent can be caused by the dissemination of droplet nuclei (aerosols) that remain infectious when suspended in the air over long distances and time. Given its potential role in the transmission of SARs-CoV-2, we recently carried out a systematic review to identify, appraise and summarise the evidence to date. [1] This review included 67 primary studies and 22 reviews reporting on airborne transmission.

All of the included studies were observational, very low-to low-quality, and lacked standardised methods. Ten studies attempted viral culture: the lack of recoverable SARS-CoV-2 virus in cultured samples prevented firm conclusions from being drawn. Other reviews suggested there was insufficient evidence to support airborne transmission. [2, 3] The majority of primary studies in these reviews detected SARS-CoV-2 RNA in the air, but not infectious virus, preventing inference of transmission from the air.

A recent systematic review from our group determined the infectiousness and potential contribution of asymptomatic transmission [4]. This review assessed transmission by including information on serial PCR cycle threshold (Ct) readings and/or viral culture and/or gene sequencing. This approach, along with contacting all authors for additional detail, permitted a more robust assessment of the evidence: the findings from higher-quality studies provided probable evidence of SARS-CoV-2 transmission from presymptomatic and asymptomatic individuals. [4]

Since the publication of our first review on airborne transmission [1], several new studies that include infectious virus status have been published. We have also proposed a framework to determine higher-quality evidence for the transmission of respiratory pathogens. [5] We therefore plan to systematically review the evidence for airborne transmission with high-level confirmatory studies of infection that include viral culture or longitudinal serial PCRs with or without gene sequencing. To assess the transmission potential of airborne transmission of SARS-CoV-2, we aim to address the following questions:

1. Are airborne samples infectious?
2. If so, what proportion are infectious, and what is the distance and duration of infectiousness in the air?
3. What is the relationship between infectiousness and airborne PCR cycle threshold (Ct)?
4. Is there evidence of a chain of transmission that establishes an actual instance of airborne transmission of SARS-CoV-2?
5. What circumstances might facilitate infectious viruses being airborne over long distances?

## Methods

We aim to identify, appraise, and summarise the evidence relating to the role of airborne transmission of SARS-CoV-2 and its relationship with infectiousness (viral culture and/or serial qRT-PCRs with or without gene sequencing) and the factors influencing transmissibility.

### Search Strategy

We will update our searches using four databases: LitCovid, medRxiv, Google Scholar, and the WHO Covid-19 database, using the terms aerosol OR airborne OR airborne OR inhalation OR air OR droplet and viral replication, viral culture, viral transmission and various synonyms from 10 September 2020 (the date our last systematic review search finished) to 30 December 2021. An information specialist [JB] will undertake the searches, and for relevant articles, will undertake citation screening to identify relevant studies (see Appendix A). Two reviewers (CJH, EAS) will independently screen study abstracts to determine eligibility. Any disagreements will be resolved through discussion. Where an agreement cannot be reached, a third reviewer (TJ) will arbitrate.

We will only include studies reporting airborne transmission attempting viral culture or serial qRT-PCR with or without genomic sequencing. We have previously defined viral culture as one of several methods to detect virus growth in cell culture in combination with a method that can identify the replicating agent as SARS-CoV-2. [4] Most commonly, this is a plaque assay combined with RT-PCR diagnosis or immunological staining or viral RNA gene sequencing (a process that helps to determine the order, or sequence, of the nucleotides in each of the genes present in a virus’s genome). Predictive or modelling studies will be excluded.

### Data extraction

We will extract variables using a standardised data extraction and assessment template. We will extract information on the study characteristics, population, setting including the environment and methods, and the main results from included studies. We will also extract data on the type of study, setting, the sampling methods including quantitative distance measures of the sampling from the source, RT-PCR samples for SARS-CoV-2 RNA including cycle threshold (Ct) and copies per m^3^ or other quantitative measures of sampled air, viral culture methods and results, size of air particles (when reported) and proportion in the sample.

Search yields will be screened in duplicate and data from included studies will be extracted into templates including study characteristics, methodological aspects of studies and a summary of the main findings. Two reviewers (CJH, EAS) will independently extract data on the inclusion criteria for the review and data for the transmission analysis. One reviewer will extract data for each study, and a second reviewer will check the extraction. We will tabulate data and summarise the data descriptively and report the duration of detectable RNA and the relationship of PCR cycle threshold and log 10 copies to positive viral culture.

To assess the chain of transmission (question 4), we will use methods based on our previously published review on presymptomatic and asymptomatic individuals [4] that only include studies with

a. documentation of the likelihood of transmission;
b. presence of infectious virus from viral culture (defined as encompassing any of several methods whereby one can detect exponential virus growth in cell culture in combination with a method that can uniquely identify the replicating agent as being SARS-CoV-2) and/or documentation of phylogenetics (i.e., genetic sequence lineage); and/or
c. adequate follow-up and reporting of symptoms and signs [6, 7]

For explanations and appendices see supplementary material, on figshare. Dataset Available at https://doi.org/10.6084/m9.figshare.18332996.v1

Appendix A: Search Strategy

Appendix B: Transmission Assessment Explainer

Appendix C: Standardised transmission of SARS-CoV-2 case causality assessment

### Quality Assessment

We have previously published methods for assessing quality [1, 4] that required adaptation based on the mode of transmission. To assess quality we will judge the following domains:

i. Source population – was the population recruited into the study clearly described?
ii. Methods – did the study authors sufficiently describe the methods used to enable replication of the study?
iii. Sample sources – were sources for the air samples clear?
iv. Outcome reporting – was the analysis of the results appropriate, and
v. Follow-up – was the pattern and number of air samples sufficient to demonstrate airborne transmission.

Where necessary, one reviewer will contact the corresponding authors of the included papers for additional information and include the responses in our assessment of bias. One reviewer (EAS) will categorize the potential for bias as high, moderate, or low which will be independently checked by a second reviewer (CJH). Reasons for the bias assessment for each study will also be recorded.

Disagreements will be resolved through discussion with the help of a third reviewer (TJ).

### Data Analysis

We will follow PRISMA reporting guidelines for systematic reviews [8]. We will evaluate the overall association between airborne transmission and SARS-CoV-2 culture with the results of qRT-PCR and variables that may influence the interpretation of the test, such as time from symptom onset and patient factors. Where the data permits we will undertake meta-analyses using R and Stata software packages. Where feasible, we will perform subgroup analyses to assess outcomes by environmental setting (e.g., indoors or outdoors).

## Data Availability

All data included in the review and the Appendices, tables and text will be made available at Figshare.

https://doi.org/10.6084/m9.figshare.18332996.v1

## Ethics committee approval

No ethics approval is necessary.

## Data Availability

All data included in the review and the Appendices, tables and text will be made available at Figshare: https://doi.org/10.6084/m9.figshare.18332996.v1 files available to download.

## Conflict of interest statements

TJ received a Cochrane Methods Innovations Fund grant to develop guidance on using regulatory data in Cochrane reviews (2015 to 2018). From 2014 to 2016, he was a member of three advisory boards for Boehringer Ingelheim. TJ was a member of an independent data monitoring committee for a Sanofi Pasteur clinical trial on an influenza vaccine. Market research companies occasionally interview TJ about phase I or II pharmaceutical products for which he receives fees (current). TJ was a member of three advisory boards for Boehringer Ingelheim (2014 to 16). TJ was a member of an independent data monitoring committee for a Sanofi Pasteur clinical trial on an influenza vaccine (2015 to 2017). TJ is a relator in a False Claims Act lawsuit on behalf of the United States that involves sales of Tamiflu for pandemic stockpiling. If resolved in the United States favour, he would be entitled to a percentage of the recovery. TJ is coholder of a Laura and John Arnold Foundation grant for the development of a RIAT support centre (2017 to 2020) and Jean Monnet Network Grant, 2017 to 2020 for The Jean Monnet Health Law and Policy Network. TJ is an unpaid collaborator to the Beyond Transparency in Pharmaceutical Research and Regulation led by Dalhousie University and funded by the Canadian Institutes of Health Research (2018 to 2022). TJ consulted for Illumina LLC on next-generation gene sequencing (2019 to 2020). TJ was the consultant scientific coordinator for the HTA Medical Technology programme of the Agenzia per I Servizi Sanitari Nazionali (AGENAS) of the Italian MoH (2007 to 2019). TJ is Director Medical Affairs for BC Solutions, a market access company for medical devices in Europe. TJ was funded by NIHR UK and the World Health Organization (WHO) to update Cochrane review A122, Physical Interventions to interrupt the spread of respiratory viruses. Oxford University funds TJ to carry out a living review on the transmission epidemiology of COVID 19. Since 2020, TJ receives fees for articles published by The Spectator and other media outlets. TJ is part of a review group carrying out a Living rapid literature review on the modes of transmission of SARS CoV 2 (WHO Registration 2020/1077093 0). He is a member of the WHO COVID 19 Infection Prevention and Control Research Working Group, for which he receives no funds. TJ is funded to co-author rapid reviews on the impact of Covid restrictions by the Collateral Global Organisation.

CJH holds grant funding from the NIHR, the NIHR School of Primary Care Research, the NIHR BRC Oxford and the World Health Organization for a series of Living rapid reviews on the modes of transmission of SARs CoV 2, reference WHO registration No2020/1077093, and to carry out a scoping review of systematic reviews of interventions to improve vaccination uptake, reference WHO Registration 2021/1138353-0. He has received financial remuneration from an asbestos case and given legal advice on mesh and hormone pregnancy tests cases. He has received expenses and fees for his media work, including occasional payments from BBC Radio 4 Inside Health and The Spectator. He receives expenses for teaching EBM and is also paid for his GP work in NHS out of hours (contract Oxford Health NHS Foundation Trust). He has also received income from the publication of a series of toolkit books and appraising treatment recommendations in non-NHS settings. He is the Director of CEBM, an NIHR Senior Investigator and an advisor to Collateral Global.

DHE holds grant funding from the Canadian Institutes for Health Research and Li Ka Shing Institute of Virology relating to the development of Covid 19 vaccines and the Canadian Natural Science and Engineering Research Council concerning Covid 19 aerosol transmission. He is a recipient of World Health Organization and Province of Alberta funding which supports the provision of BSL3 based SARS CoV 2 culture services to regional investigators. He also holds public and private sector contract funding relating to the development of poxvirus based Covid 19 vaccines, SARS CoV 2 inactivation technologies, and serum neutralization testing.

JMC holds grants from the Canadian Institutes for Health Research on acute and primary care preparedness for COVID 19 in Alberta, Canada and was the primary local Investigator for a *Staphylococcus aureus* vaccine study funded by Pfizer, for which all funding was provided only to the University of Calgary. He is a co-investigator on a WHO funded study using integrated human factors and ethnography approaches to identify and scale innovative IPC guidance implementation supports in primary care with a focus on low resource settings and using drone aerial systems to deliver medical supplies and PPE to remote First Nations communities during the COVID 19 pandemic. He also received support from the Centers for Disease Control and Prevention (CDC) to attend an Infection Control Think Tank Meeting. He is a member and Chair of the WHO Infection Prevention and Control Research and Development Expert Group for COVID 19 and the WHO Health Emergencies Programme (WHE) Ad hoc COVID 19 IPC Guidance Development Group, both of which provide multidisciplinary advice to the WHO, for which no funding is received and from which no funding recommendations are made for any WHO contracts or grants. He is also a member of the Cochrane Acute Respiratory Infections Group.

JB is a major shareholder in the Trip Database search engine (www.tripdatabase.com) as well as being an employee. In relation to this work, Trip has worked with a large number of organizations over the years; none have any links with this work. The main current projects are with AXA and Collateral Global.

ECR was a member of the European Federation of Neurological Societies(EFNS) / European Academy of Neurology (EAN) Scientist Panel, Subcommittee of Infectious Diseases (2013 to 2017). Since 2021, she is a member of the International Parkinson and Movement Disorder Society (MDS) Multiple System Atrophy Study Group, the Mild Cognitive Impairment in Parkinson Disease Study Group, and the Infection Related Movement Disorders Study Group. She was an External Expert and sometimes Rapporteur for COST proposals (2013, 2016, 2017, 2018, 2019) for Neurology projects. She is a Scientific Officer for the Romanian National Council for Scientific Research.

SM is a pharmacist working for the Italian National Health System since 2002 and a member of one of the three Institutional Review Boards of Emilia-Romagna Region (Comitato Etico Area Vasta Emilia Centro) since 2018.

AP holds grant funding from the NIHR School of Primary Care Research. IJO and EAS have no interests to disclose.

